# Growth Trajectories in Ascending Thoracic Aortic Dilation: Classification and Implications for Imaging Surveillance

**DOI:** 10.1101/2025.06.27.25330454

**Authors:** Prabhvir S. Marway, Carlos Alberto Campello Jorge, Gregory Spahlinger, Marion Hofmann-Bowman, Matthew S. Davenport, Nicholas S. Burris

## Abstract

**Background:** Ascending aortic dilation is monitored with serial imaging, yet event rates are very low, with most type A dissections occurring at non-surgical sizes limiting their prevention by prophylactic repair. We aimed to characterize ascending aortic growth trajectories in a real-world population to understand their clinical consequences and better inform surveillance strategies.

**Methods and Results:** We retrospectively studied 3,363 adults (median age 62 years; 68 % men) with ≥2 chest CTA/MRA examinations at a single center. Mid-ascending diameters were extracted from structured reports and clustered with latent profile analysis (LPA). Early (first 3 scans; n= 1,997) and Extended (first 5 scans; n= 757) models each yielded four classes — Stable, Growth, Dramatic Growth and Non-physiologic. The Stable class was most common across follow-up (74 % Early; 70 % Extended); Growth and Dramatic Growth classes comprised 23% and 2% respectively. Only 1.1 % ever met guideline growth-based criteria for repair, and this small subgroup showed smaller baseline diameters (37.0 mm vs. 40.3 mm, p=0.009). Among patients with Extended follow-up, 80% of those initially Stable remained Stable. Reclassification into a Growth group associated with younger and age and history of Marfan syndrome (50.0% vs 3.0%, p=0.006). Acute type A dissection was rare (0.45 %) and not clearly linked to any trajectory.

**Conclusions:** Most patients with a dilated ascending aorta show negligible growth and very low complication rates. Repeated imaging beyond 3 scans may amplify uncertainty without clear improvements in risk stratification, suggesting that imaging intervals may be safely de-escalated for initially stable patients.

## Introduction

Ascending thoracic aortic aneurysms (aTAA) are managed primarily through imaging surveillance, in which aortic size is monitored in order to predict disease severity and potentially prevent life-threatening complications such as type A aortic dissection (TAAD). Current guidelines recommend imaging surveillance for ascending aortic diameters >40 mm,^1^ with approximately 7 million adults in the US meeting this criteria.^2–4^ A diameter of ≥55mm is a Class I indication for prophylactic surgical repair of the ascending aorta.^1^ However, aneurysms meeting surgical size criteria are infrequent even in the clinical surveillance population (∼4%).^5^ While the risk of aortic complications such as TAAD does increase with aortic size,^5–7^ the vast majority (>90%) of dissections occur in patients well below surgical repair thresholds,^8^ and often barely within surveillance size ranges (median pre-dissection diameter of ∼40mm).^9^ This apparent ‘paradox’ has been explained by the incidence of large aneurysms being very rare in comparison to mild degrees of aortic dilation, despite their higher risk.

In addition to absolute size, the growth rate of aTAA has been proposed as an independent risk factor for TAAD, with indirect evidence suggesting a 2-3-fold increased risk of TAAD in aTAAs that are growing compared to aTAAs that are not.^5^ However, there is no clear evidence as to what specific growth rates confer what risk. Recent studies in aTAA surveillance populations have shown mean growth rates of only 0.3 mm/year in the asymptomatic aTAA surveillance population,^10,11^ a growth rate which is challenging to measure reliably over typical annual or biennial surveillance intervals. Recently revised guidelines have proposed growth of ≥ 5 mm in one year, or ≥ 3mm/year in two consecutive years, as indications for prophylactic surgical repair. However, these thresholds have been largely informed by the levels of change in aortic size that can be confidently detected through manual measurements, which are known to be imprecise (i.e., variability of up to 3-5 mm).^12^ In addition, recent reports have emphasized that the occurrence of “real” aortic growth above guideline thresholds is rare.^13,14^ All this calls into question the clinical utility of long-term imaging surveillance in aTAA, especially in patients who are far from absolute diameter repair thresholds.

One key obstacle to prescribing optimal imaging-surveillance strategies is our limited understanding of the distribution of growth trajectories in the ascending aorta. Current approaches largely rely on average rates, potentially masking distinct subgroups with unique risk profiles. Consequently, optimizing imaging utilization and surveillance frequency—as well as identifying the patient characteristics that should guide monitoring intensity—remain significant challenges. Recognizing that aortic dissection commonly occurs at diameters below conventional surgical thresholds and aiming to more fully capture the natural history of growth patterns during surveillance, this study aimed to include all patients undergoing serial, cross-sectional imaging for surveillance of vascular disease, including standardized measurements of the ascending aorta, regardless of baseline aortic size. We sought to apply automated clustering techniques, specifically a technique termed latent profile analysis (LPA), to track longitudinal changes in aortic diameter in a large, real-world clinical cohort to identify sub-groups with differing growth patterns. Our objectives were to: (1) identify distinct aortic growth trajectories classes and determine their prevalence; (2) investigate associations between trajectory class membership, patient characteristics and subsequent clinical outcomes (TAAD, prophylactic repair); and (3) evaluate the stability of growth trajectory class membership over extended surveillance to better understand the clinical utility of life-long surveillance paradigms.

## Methods

### Study Population and Data Collection

We included all adult patients with 2 or more CT or MR angiograms (CTA or MRA) of the chest with full coverage of a native ascending aorta (i.e., unrepaired and without acute aortic pathology), with >6 months of total surveillance at a single University health system, imaged between January 2000 and December 2023. Ethical approval was obtained via Institutional Review Board review (HUM00133798). Imaging studies within 3 months of each other were not considered as distinct measurements, with the largest measurement values taken in such cases. Patients were initially identified through a clinical programmatic database of aortic patients, with all cases undergoing subsequent review of imaging and clinical charts to ensure pre-existing aortic repairs or chronic dissections were excluded, and to collect baseline demographics, comorbidities (e.g., hypertension, diabetes mellitus, smoking history, known heritable thoracic aortic disease and confirmed chart diagnosis of aortitis), vital signs (e.g., blood pressure), medications, and clinical outcomes (e.g., type A aortic dissection, prophylactic aortic repair, aorta-specific mortality).

### Aortic Diameter Measurement

All clinical aortic measurements at our institution are performed by a dedicated 3D lab staffed by trained technicians, employing standardized measurement protocols and centerline analysis, and are verified by cardiothoracic radiologists. Aortic diameter data was extracted from templated clinical imaging reports by an in-house text processing pipeline that searches for pre-specified keywords for each aortic landmark and identifies associated diameter values. To ensure consistency of measurement location across all cases and time-points, the mid-ascending aorta (mid-AsAo) landmark was used to extract diameter measurements for analysis, with manual review performed in cases with suspected typographical errors for implausible values (e.g., interval change >10 mm, absolute size >70mm or <20mm).

### Latent profile analysis (LPA)

To identify subgroups of patients with distinct aortic growth patterns, we used latent profile analysis (LPA), a statistical clustering technique based in Gaussian finite mixture modelling, to that empirically classify patients based on their longitudinal diameter changes without requiring pre-specified growth thresholds.

Separate LPA models were developed to assess growth trajectories at different surveillance durations. An “Early” classification was determined using each patient’s first 3 scans, as three timepoints are the minimum required to establish a growth trend. An “Extended” classification was determined using the first 5 scans to assess longer-term trajectory stability. The resulting classes were analyzed to determine their prevalence, their association with patient characteristics and clinical outcomes, and the rate at which patients transitioned between classes over time. Full details of the LPA methodology are provided in the **Supplemental Methods**.

### Statistical analysis

Continuous variables were summarized as medians with interquartile ranges (IQR), and categorical variables as counts and percentages. Comparisons between groups (e.g., based on number of scans, LPA classes) were performed using the Kruskal-Wallis test for continuous variables and the Chi-squared test or Fisher’s exact test (for cell counts <5) for categorical variables. Pairwise comparisons were conducted using appropriate post-hoc tests if the overall test was significant (p < 0.05). All statistical analyses were performed using Python (version 3.13). This study was conducted and reported in accordance with the Strengthening the Reporting of Observational Studies in Epidemiology (STROBE) guidelines.

## Results

### Study population

Overall, we identified 3.363 patients with 2 or more CTAs and/or MRAs meeting inclusion criteria. Patients had a median age of 62.3 (IQR: 53.1, 70.7) years, 68% (n=2287) were male, and 84% (n=2842) had a history of hypertension. Heritable thoracic aortic disease was present in a minority (3.4%, n=115), with Marfan syndrome being the most common (2.6%, n=86). A modest number (10%, n=337) of patients underwent eventual prophylactic ascending aorta repair and 0.45% (n=15) experienced acute TAAD. The overall cohort median baseline mid-AsAo diameter was 40.2 mm (IQR: 36.0, 44.0), and the median growth rate was 0.079 mm/year (IQR: -0.134, 0.370) over a median total imaging surveillance period of 3.50 years (IQR: 1.60, 6.42). Approximately a quarter of patients (28%, n=946) had concomitant aortic pathology involving arch, descending, or abdominal aortic segments, **Table 1**.

**Table 1:**
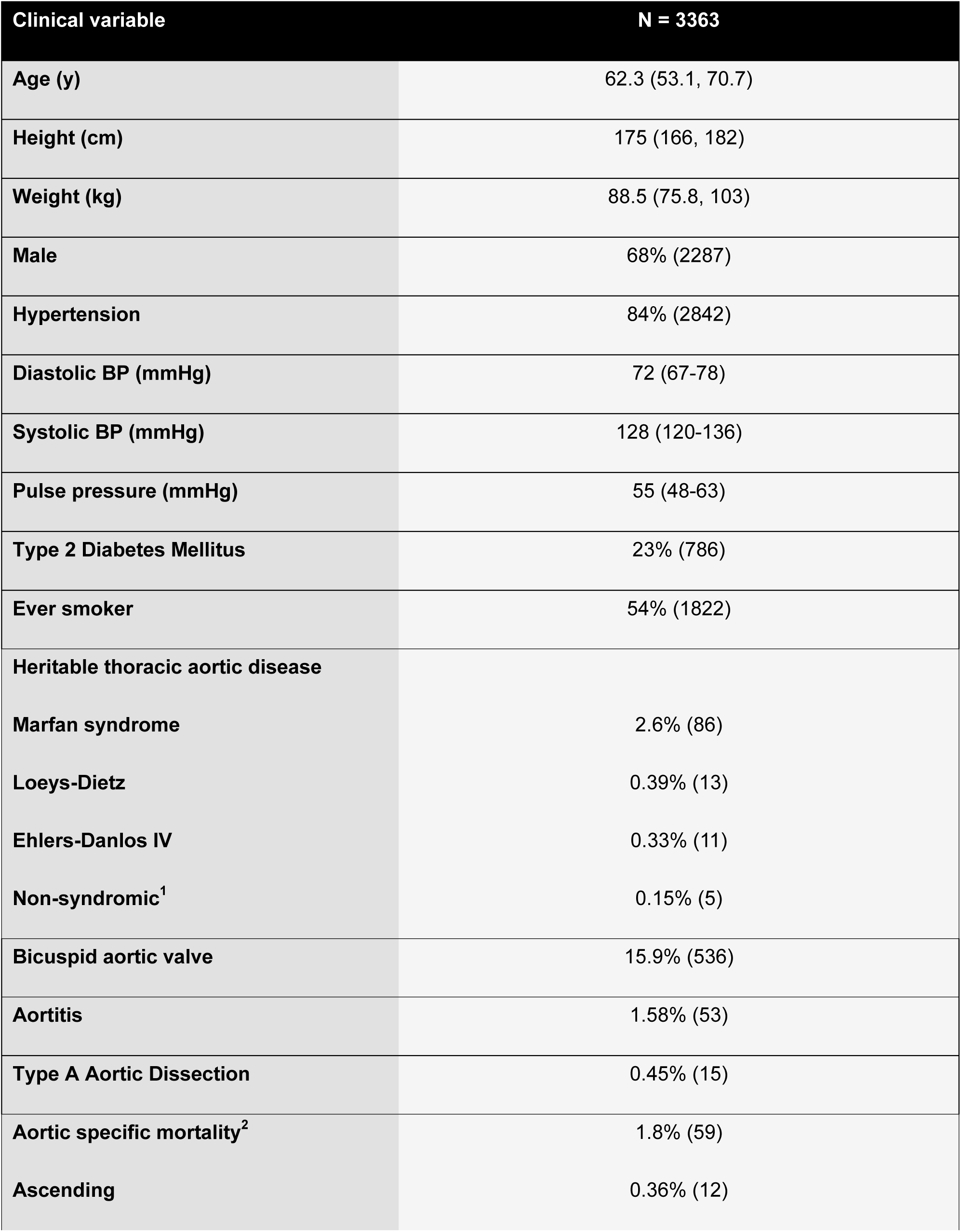
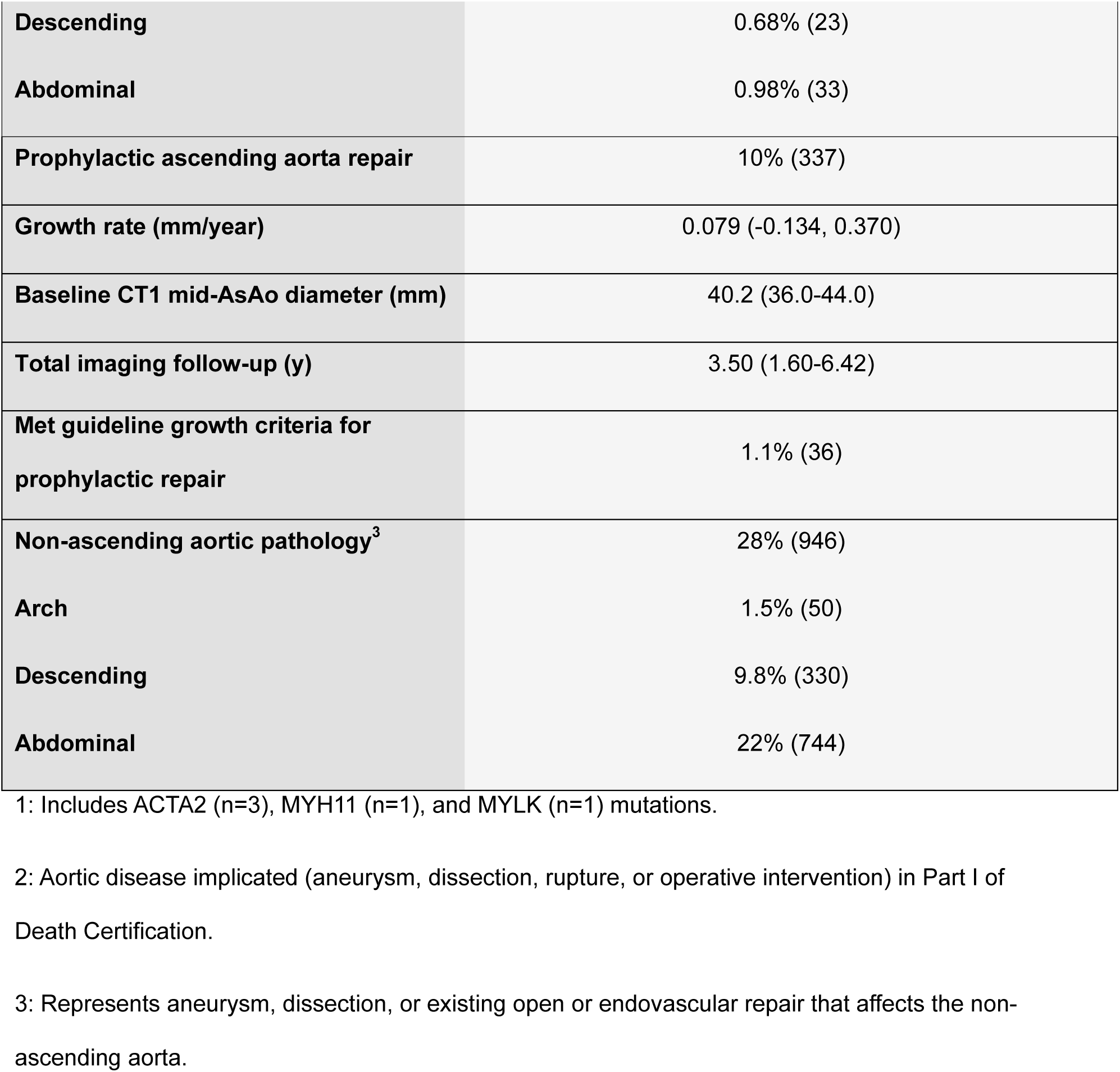
Overall population demographics and outcomes.

| Clinical variable | N = 3363 |
| --- | --- |
| Age (y) | 62.3 (53.1, 70.7) |
| Height (cm) | 175 (166, 182) |
| Weight (kg) | 88.5 (75.8, 103) |
| Male | 68% (2287) |
| Hypertension | 84% (2842) |
| Diastolic BP (mmHg) | 72 (67-78) |
| Systolic BP (mmHg) | 128 (120-136) |
| Pulse pressure (mmHg) | 55 (48-63) |
| Type 2 Diabetes Mellitus | 23% (786) |
| Ever smoker | 54% (1822) |
| Heritable thoracic aortic disease |  |
| Marfan syndrome | 2.6% (86) |
| Loeys-Dietz | 0.39% (13) |
| Ehlers-Danlos IV | 0.33% (11) |
| Non-syndromic<sup>1</sup> | 0.15% (5) |
| Bicuspid aortic valve | 15.9% (536) |
| Aortitis | 1.58% (53) |
| Type A Aortic Dissection | 0.45% (15) |
| Aortic specific mortality<sup>2</sup> | 1.8% (59) |
| Ascending | 0.36% (12) |
| **Descending** | 0.68% (23) |
| **Abdominal** | 0.98% (33) |
| **Prophylactic ascending aorta repair** | 10% (337) |
| **Growth rate (mm/year)** | 0.079 (-0.134, 0.370) |
| **Baseline CT1 mid-AsAo diameter (mm)** | 40.2 (36.0-44.0) |
| **Total imaging follow-up (y)** | 3.50 (1.60-6.42) |
| **Met guideline growth criteria for prophylactic repair** | 1.1% (36) |
| **Non-ascending aortic pathology<sup>3</sup>** | 28% (946) |
| **Arch** | 1.5% (50) |
| **Descending** | 9.8% (330) |
| **Abdominal** | 22% (744) |

1: Includes ACTA2 (n=3), MYH11 (n=1), and MYLK (n=1) mutations.
2: Aortic disease implicated (aneurysm, dissection, rupture, or operative intervention) in Part I of Death Certification.
3: Represents aneurysm, dissection, or existing open or endovascular repair that affects the non-ascending aorta.

**Supplemental Table 1** stratifies demographics and outcomes by the number of scans of the native ascending aorta available per patient (2, 3, 4, or 5+). Compared to patients with 2 available scans, those with ≥5 scans were slightly younger (median 60.2 vs. 63.3 years, p<0.001), had higher rates of hypertension (90.0% vs 80.0%, p<0.001) and Marfan syndrome (4.1% vs 1.8%, p=0.013), experienced TAAD more frequently (0.786% vs 0.073%, p=0.004), exhibited a slightly higher median growth rate (0.13 mm/year vs 0.00 mm/year, p<0.001), and were more likely to have concomitant arch, descending, or abdominal aortic pathology (39% vs 23%, p<0.001).

### Guideline Growth Thresholds

Based on current guidelines, only 1.1% (n=36) of patients met growth criteria for repair at any point in their imaging surveillance, with 34 patients growing ≥ 5mm within 1 year (or ≥ 5 mm/year annualized over at least 1 year) and 17 growing ≥ 3 mm/year annualized over at least 2 years. Compared to those who never met growth-based criteria for repair, these patients were more likely to have smaller baseline mid-AsAo diameters (37.0 mm [IQR: 31.8, 41.9] vs. 40.3 mm [IQR: 36.0, 44.0], p=0.009), have more surveillance imaging (5 scans [IQR: 3, 6] vs. 3 scans [IQR: 2, 4], p<0.001), have Marfan syndrome (11.1% vs. 2.5%, p=0.013), and have higher rates of arch, descending, or abdominal aortic pathology (44.4% vs. 23.1%, p<0.001). Rates of acute TAAD during surveillance (0% vs 0.45%, p>0.99), prophylactic repair (13.9% vs 10.0%, p= 0.618), and all other demographic and clinical factors were not significantly different between patients who did vs. did not meet growth criteria for repair (p>0.05).

### Latent Profile Analysis (LPA)

LPA was performed on the subset of patients with at least 3 scans of the native ascending aorta (n=1997) to determine an “Early” growth classification. Using each patient’s first 3 scans, a 4-class classification scheme was statistically derived based on optimization metrics (BIC) and preservation of clinically meaningful groups. The resulting classes were given names that were felt to best describe their semantic interpretation, with the 4 classes being: Stable (Class 1), Growth (Class 2), Dramatic Growth (Class 3), and Contraction/Non-physiologic (Class 4) given that true aortic growth is assumed to occur by monotonic increases in size rather than bidirectional changes. Similarly, LPA analysis was also performed on those patients with at least 5 scans of the native ascending aorta (n=757) to determine an “Extended” growth classification. A similar 4-class semantic interpretation was also optimal for this subgroup, with analogous classes to the “Early” growth classification (3-scan) identified: Stable (Class 1), Growth (Class 2), Dramatic Growth (Class 3), and Non-physiologic (Class 4). **Figure 1** depicts the growth trajectories for all patients as stratified by the “Extended” LPA growth classification (5 scans).

**Figure 1:**
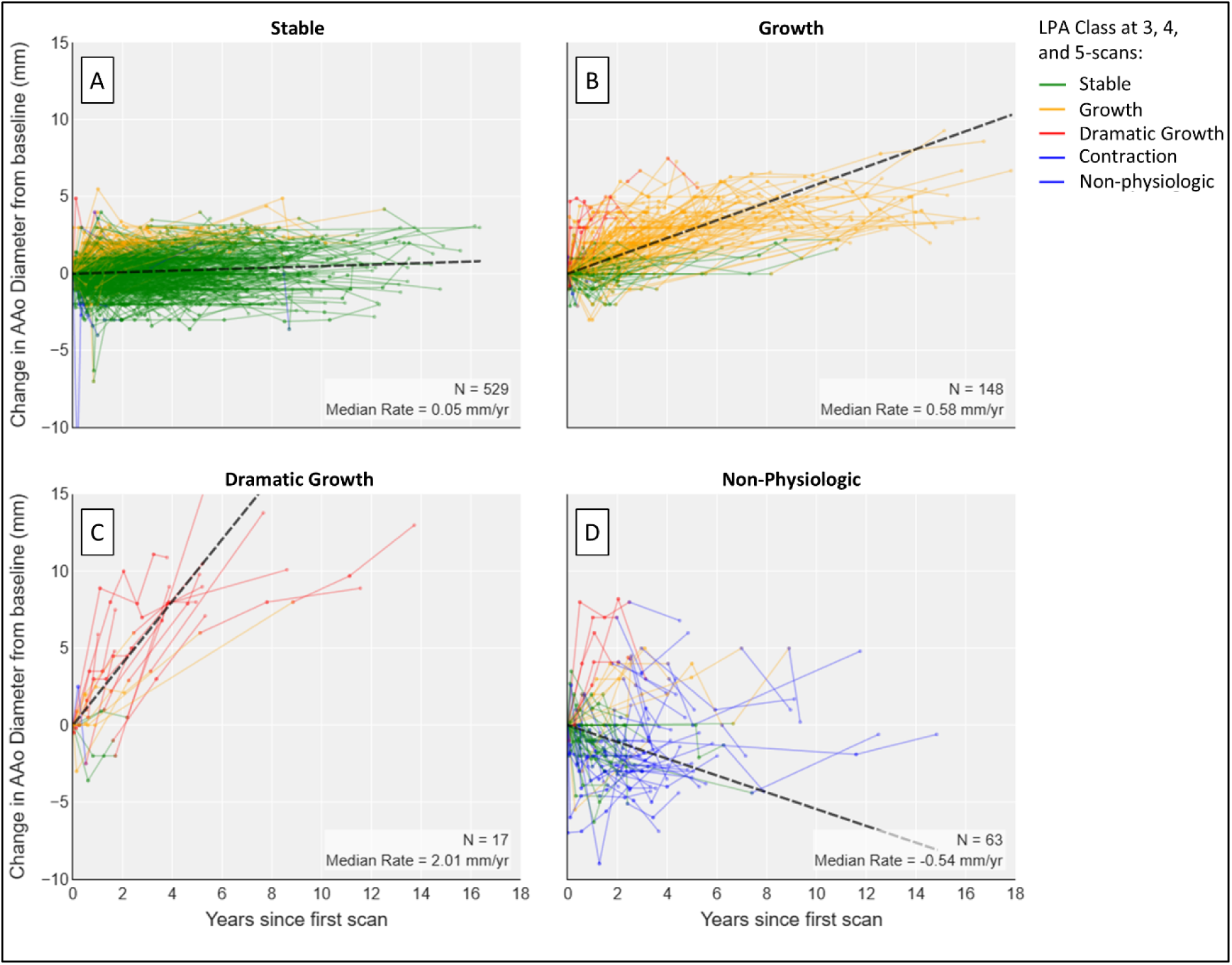
Growth trajectories for all patients in the 5-scan LPA analysis (N=757), stratified by classification (**A**: Stable, **B**: Growth, **C**: Dramatic Growth, **D**: Non-Physiologic) at the 5-scan LPA. Trajectory lines are colorized based on the LPA class assigned to the first 3, 4, and 5 scans for each patient, leading to line color changing for patients who experienced class cross-over between 3 and 5 scans.

In both the “Early” and “Extended” LPA growth classification, the Stable groups were by far the largest (74.2% and 69.6% respectively), with smaller representations in the “Growth” (22.6% and 19.6%) and “Dramatic Growth” (1.85% and 2.25%) groups. As expected, LPA-derived growth rates differed significantly between classes (p<0.001 for both “Early” and “Extended” growth classification) with overall growth rates showing a decreasing trend over time (i.e., slowing growth) for both the Growth and Dramatic Growth groups, p<0.001 (**Table 2**).

**Table 2:**
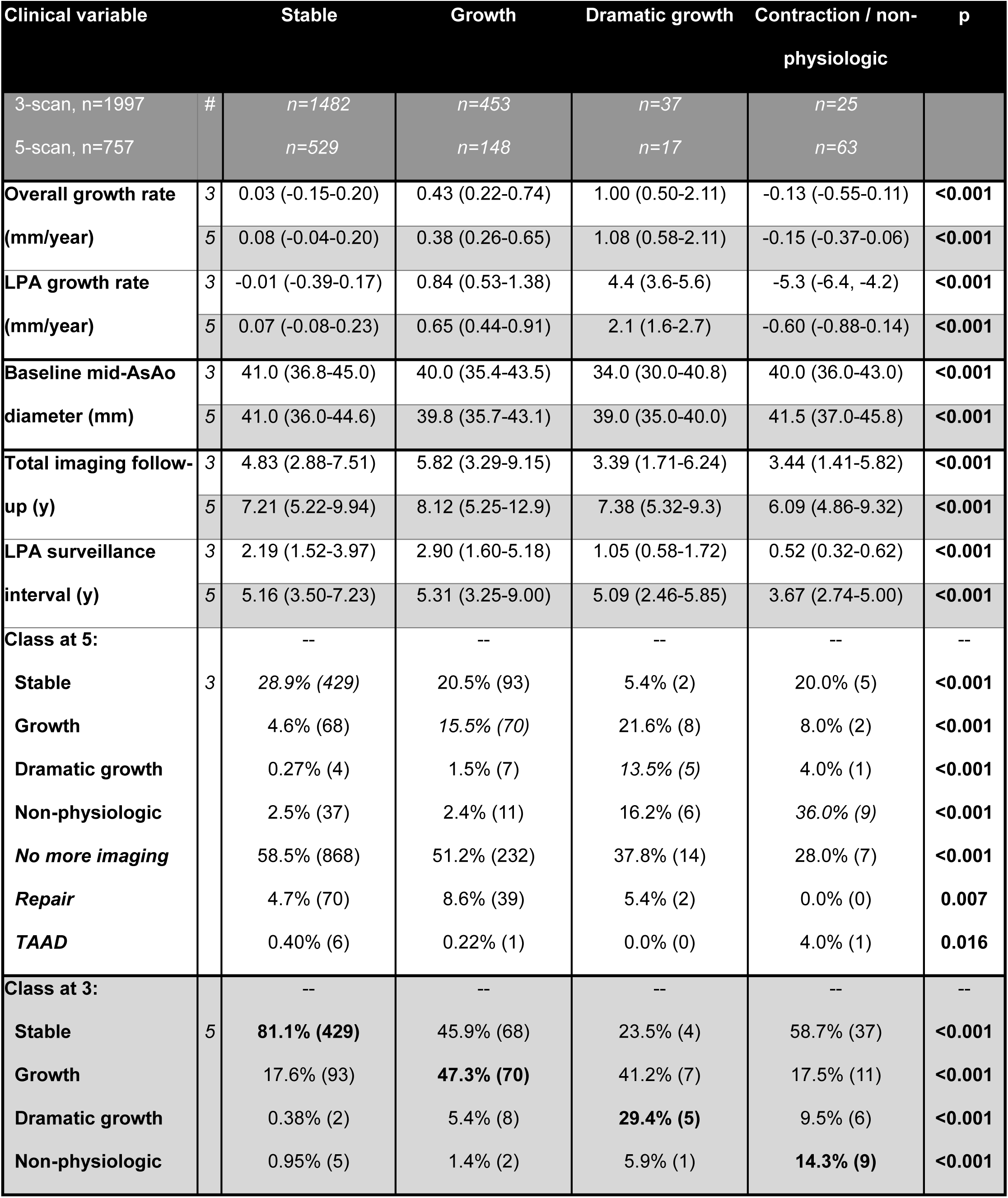
Growth and group cross-over data stratified by LPA class using first 3 (“Early”) and the first 5 (“Extended”) imaging studies. Values given as median (IQR) or frequency % (n).

### “Early” LPA Growth Class (3-Scan) Comparisons

In 3-scan analysis, the median baseline mid-AsAo diameter was significantly lower in the Dramatic Growth class vs. the Stable class (34.0 mm vs 41.0 mm, p<0.001), **Table 2**. Demographic data and rates of Marfan syndrome did not differ significantly across latent classes. Bicuspid aortic valve disease was most frequently seen in the Growth group (21.2%), and least likely in the Dramatic Growth (2.7%) group (p<0.001). Prophylactic ascending aorta repair rates increased significantly from Non-physiologic (0%) to Stable (8.2%) to Growth (14.6%) groups but were intermediate in the small group (n=37) of Dramatic Growth (10.8%, overall p<0.001). Acute TAAD rates were highest in the Non-physiologic (8.0%) and Dramatic Growth (2.7%) groups compared to Stable (0.61%) and Growth (0.4%), overall p<0.001. The frequency of arch, descending, or abdominal aortic pathology was significantly higher in Dramatic Growth (46%) compared to Stable (30%), p<0.001. Ascending aorta-specific mortality was higher in the Growth (4.1%) and Dramatic Growth (5.9%) compared to Stable (1.5%), but this did not reach statistical significance (p=0.182) (**Table 3**).

**Table 3:** Clinical demographics and outcomes stratified by LPA class using first 3 (“Early”) and the first 5 (“Extended”) imaging studies. Values given as mean (SD), median (IQR) or frequency % (n). 1: Aortic disease implicated (aneurysm, dissection, rupture, or operative intervention) in Part I of Death Certification. 2: Represents aneurysm, dissection, or existing open or endovascular repair that affects the non-ascending aorta.

| Clinical variables |  | Stable | Growth | Dramatic growth | Contraction / non-physiologic | p |
| --- | --- | --- | --- | --- | --- | --- |
| 3-scan, n=1997 |  | n=1482 | n=453 | n=37 | n=25 |  |
| 5-scan, n=757 |  | n=529 | n=148 | n=17 | n=63 |  |
| Age (y) | 3 | 62.1 (52.5-70.2) | 60.6 (50.6-68.3) | 59.6 (46.4-69.1) | 50.7 (45.7-63.7) | **0.004** |
|  | 5 | 61.4 (52.8-69.0) | 54.8 (46.8-65.1) | 61.8 (50.4-67.9) | 55.8 (46.7-64.6) | **<0.001** |
| Height (cm) | 3 | 175 (166-181) | 175 (168-183) | 175 (165-178) | 176 (168-183) | 0.177 |
|  | 5 | 175 (166-180) | 176 (168-183) | 170 (168-183) | 178 (168-181) | 0.278 |
| Weight (kg) | 3 | 88.4 (75.1-103) | 90.6 (77.1-106) | 86.1 (74.7-99.2) | 89.2 (80.3-107) | 0.217 |
|  | 5 | 89.8 (75.8-104) | 92.3 (79.8-105) | 79.7 (67.9-101) | 94.4 (84.8-104) | 0.098 |
| Male | 3 | 68.4% (1013) | 71.5% (323) | 56.8% (21) | 76% (19) | 0.199 |
|  | 5 | 69.2% (366) | 72.3% (107) | 52.9% (9) | 71.4% (45) | 0.410 |
| Hypertension | 3 | 88% (1297) | 87% (394) | 867% (32) | 92% (23) | 0.896 |
|  | 5 | 90.5% (479) | 87.8% (130) | 100% (15) | 93.7% (59) | 0.292 |
| Type 2 Diabetes | 3 | 23% (342) | 25% (113) | 19% (7) | 12% (3) | 0.405 |
|  | 5 | 22% (117) | 22% (32) | 12% (2) | 21% (13) | 0.781 |
| Ever smoker | 3 | 54% (804) | 53% (242) | 65% (24) | 68% (17) | 0.300 |
|  | 5 | 54.8% (290) | 58.1% (86) | 58.8% (10) | 55.6% (35) | 0.901 |
| Marfan syndrome | 3 | 2.7% (40) | 4.2% (19) | 5.4% (2) | 4.0% (1) | 0.354 |
|  | 5 | 3.2% (17) | 6.8% (10) | 12% (2) | 1.6% (1) | 0.058 |
| Bicuspid aortic valve | 3 | 15.2% (225) | 21.2% (96) | 2.7% (1) | 8.0% (2) | **0.001** |
|  | 5 | 16.1% (85) | 18.9% (28) | 5.9% (1) | 22.2% (14) | 0.330 |
| Aortitis | 3 | 1.6% (23) | 0.44% (2) | 2.7% (1) | 0.0% (0) | 0.240 |
|  | 5 | 1.3% (7) | 2.7% (4) | 12% (2) | 3.2% (2) | **0.015** |
| **Type A Aortic Dissection** | 3 | 0.61% (9) | 0.4% (2) | 2.7% (1) | 8.0% (2) | **<0.001** |
|  | 5 | 0.38% (2) | 2.0% (3) | 0.0% (0) | 1.7% (1) | 0.763 |
| **Aortic specific mortality<sup>1</sup>** | 3 | 1.5% (22) | 2.4% (11) | 5.4% (2) | 4.0% (1) | 0.244 |
|  | 5 | 1.5% (8) | 4.1% (6) | 5.9% (1) | 1.6% (1) | 0.182 |
| **Ascending** | 3 | 0.34% (5) | 0.44% (2) | 2.7% (1) | 0.0% (0) | 0.193 |
|  | 5 | 0.38% (2) | 1.4% (2) | 5.9% (1) | 0.0% (0) | **0.027** |
| **Descending** | 3 | 0.61% (9) | 1.3% (6) | 2.7% (1) | 4.0% (1) | 0.035 |
|  | 5 | 0.57% (3) | 2.7% (4) | 5.9% (1) | 1.6% (1) | **0.048** |
| **Abdominal** | 3 | 0.81% (12) | 1.3% (6) | 0.0% (0) | 0.0% (0) | 0.772 |
|  | 5 | 0.77% (4) | 1.4% (2) | 0.0% (0) | 0.0% (0) | 0.744 |
| **Prophylactic ascending aorta repair** | 3 | 8.2% (122) | 14.6% (66) | 10.8% (4) | 0.0% (0) | **<0.001** |
|  | 5 | 8.7% (46) | 16.2% (24) | 29.4% (5) | 9.5% (6) | **0.004** |
| **Non-ascending aortic pathology<sup>2</sup>** | 3 | 30% (446) | 34% (156) | 46% (17) | 32% (8) | **<0.001** |
|  | 5 | 38% (202) | 38% (56) | 47 % (8) | 40% (25) | 0.664 |

### “Extended” LPA Growth Class (5-Scan) Comparisons

In the “Extended” LPA growth subgroup, Growth and Non-Physiologic classes were significantly younger (median 54.8 years and 55.8 years, respectively) compared to other groups (overall p<0.001). Rates of Marfan syndrome were higher in the Growth (6.8%) and Dramatic Growth (12%) groups compared to Stable (3.2%) and Non-physiologic (1.6%), but this did not reach statistical significance (overall p=0.058). Aortitis rates were highest in the Dramatic Growth group (12%, overall p=0.015). Prophylactic ascending aorta repair rates were highest in the Dramatic Growth (29.4%) and Growth (16.2%) groups (overall p<0.001). Acute TAAD rates did not significantly differ between groups (p=0.762). The presence of arch, descending, or abdominal aortic pathology was significantly higher in the Dramatic Growth (47%) group compared to the Stable (38%) group (p<0.001). Ascending aorta-specific mortality significantly increased from the Stable (0.38%) to Growth (1.4%) to Dramatic Growth (5.9%) groups (overall p<0.001) (**Table 3**).

### Classification Changes Between Early and Extended Surveillance

Significant group cross-over was observed between the “Early” (3-scan) and “Extended” (5-scan) analyses, detailed in **Table 2** and visualized in **Figures 1 and 2**. Most (56.1%, 1121/1997) of the patients at the “Early” 3-scan time point did not have further imaging of the native ascending aorta, with lack of follow-up being most common in the Stable group (58.5%, 868/1482, p<0.001). Among patients with extended follow-up, those classed as Stable in the “Early” growth classification remained Stable at the “Extended” time point (79.7%, 429/538). The Dramatic Growth group at the “Early” time point was more likely to be re-classified to the Non-Physiologic (28.6%, 6/21) or Growth groups (38.1%, 8/21) than remain in the Dramatic Growth group (23.8%, 5/21) at the “Extended” time point. The Non-Physiologic group grew substantially between the “Early” and “Extended” time points (1.3% [25/1997] to 8.3% [63/757], p<0.001), with most patients (58.7%, 37/63) originating from the “Early” Stable group (**Table 2**).

**Figure 2:**
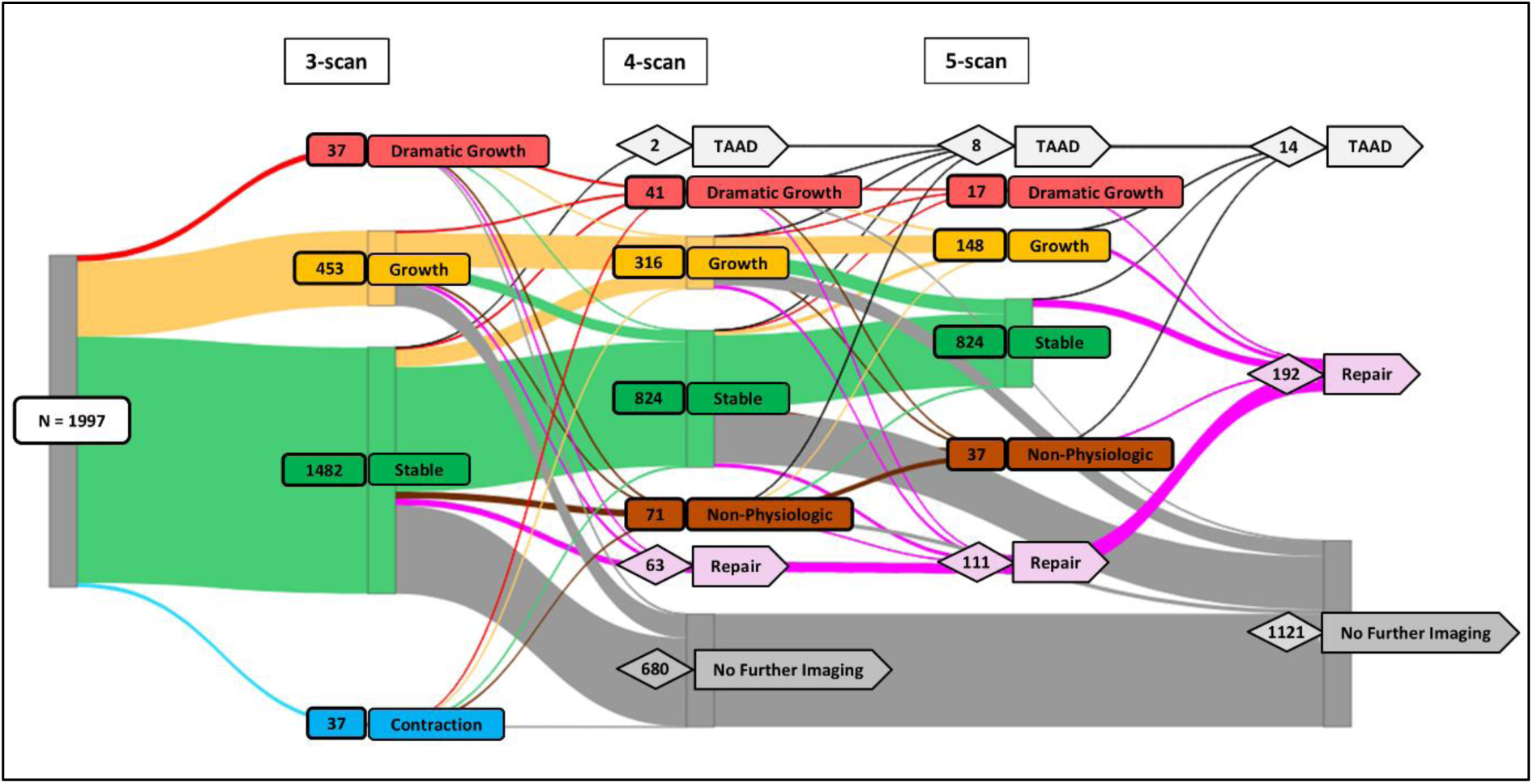
A Sankey diagram depicting how patients move between LPA classifications, and to the study endpoints, considering their first 3, 4, and 5 scans. Of note, one TAAD outcome is omitted due to lack of 3-scans. Note the extensive cross-over.

To assess which patient factors were associated with a departure from a Stable growth pattern, we stratified patients initially classified as Stable at the “Early” timepoint by their “Extended” LPA growth classification and compared baseline characteristics (**Supplemental Table 2**). Stable patients who subsequently moved into a Growth classification were younger (53.5 years [IQR: 47.6, 64.1] vs. 61.5 years [IQR: 53.2, 69.1], p<0.001), with re-classification into the Dramatic Growth associated with history of Marfan syndrome (50.0% vs 3.0%, p=0.006).

### Type A dissection subgroup

Among the small subgroup of patients that experienced type A aortic dissection during imaging surveillance (n=15), the median pre-TAAD growth rate was 0.40 mm/year (IQR: 0.11, 0.64) and median baseline mid-AsAo diameter of 40.0 mm (IQR: 38.2, 42.4). Median diameter of the mid-AsAo on the imaging study most recent to TAAD was 42.0 mm (IQR: 39.6, 43.1). **Figure 3** depicts the growth trajectory of each patient prior to TAAD, with intervals colored based on LPA classification where available. Of the 14 cases with LPA classification prior to TAAD, 50% (7) were predominantly classified Stable throughout their growth trajectory, with 14% (2) having Dramatic Growth at any point across their trajectory.

**Figure 3:**
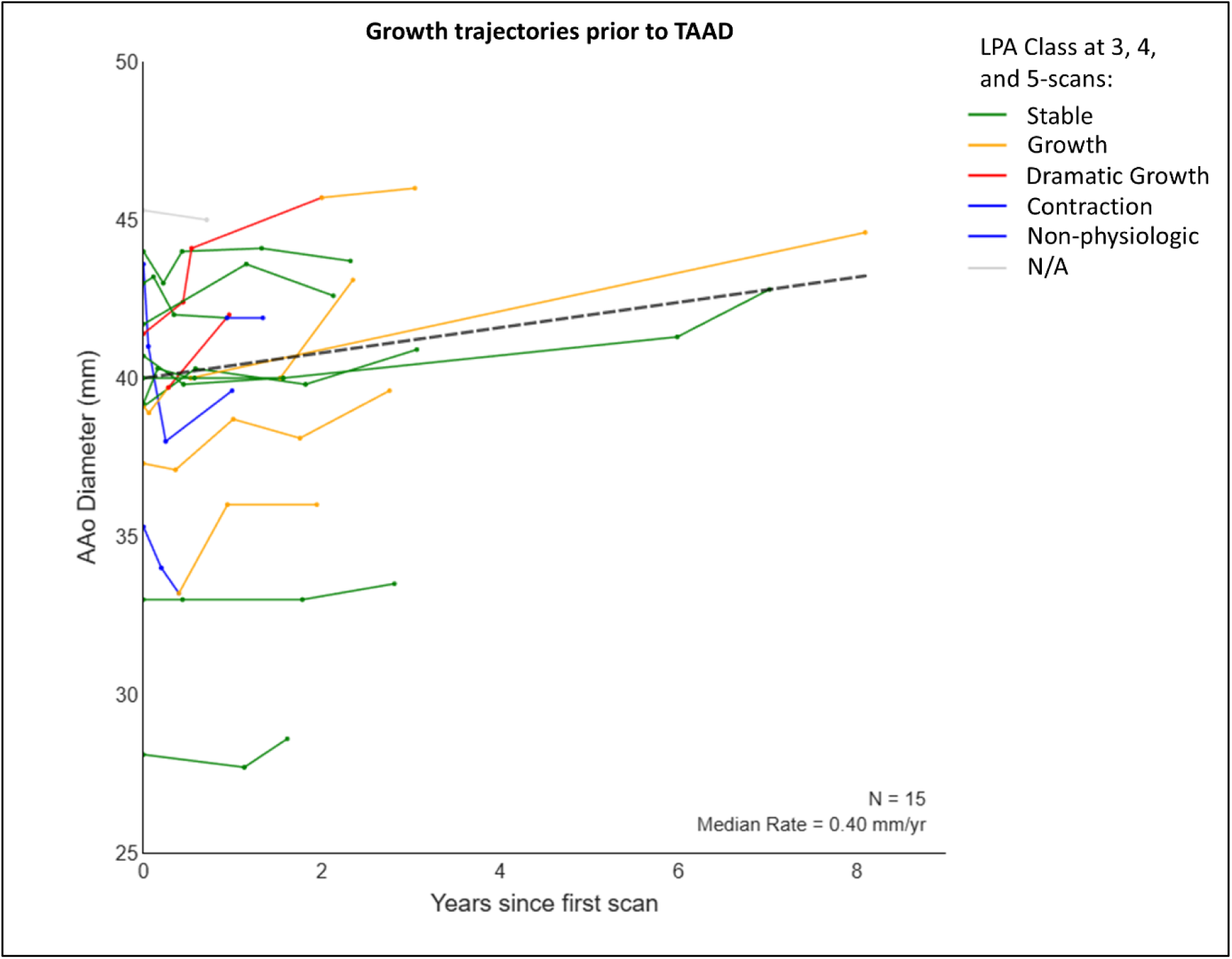
Absolute diameter growth trajectories for all patients experiencing subsequent TAAD. The first 5 diameter time-points are displayed for each patient, with trajectory lines colorized based on the calculated LPA class at the 3, 4, and 5-scan points, if applicable.

## Discussion

### Key Findings

This study examined longitudinal imaging data from a large, real-world clinical cohort of patients undergoing imaging surveillance of the ascending aorta to characterize growth trajectories and better understand the utility of current surveillance practices in aTAA. Our key findings are that most patients (>90%) exhibit stable or slow aortic growth trajectories (i.e., mean growth rates of about 0.1—0.5 mm/year) over extended follow-up (≥5 scans) when assessed with LPA, an unsupervised clustering technique that does not require pre-specified growth thresholds. Only ∼1% of patients demonstrated consistent diameter increases that could be reliably detected by annual imaging surveillance (allowing for known variability in diameter measurements) or have periods of growth rapid enough to meet the guideline criteria for repair; neither of these outcomes were clearly associated with key clinical outcomes. Importantly, when extending imaging beyond a patient’s first 3 scans, substantial cross-over between Stable and Growth groups were observed and the prevalence of Non-physiologic growth patterns increased - phenomena likely driven in part by measurement variability inherent to manual aortic measurements. As expected, patients in Growth or Dramatic Growth groups had higher rates of subsequent prophylactic repair compared to those in the Stable group. Acute TAAD was rare in our cohort (∼1 per 2000 years of patient follow-up), and although Dramatic Growth was weakly associated with TAAD compared to Stable at the “Early” time point, this association was lost with additional surveillance at the “Extended” time point.

### Comparison with existing literature

The modest median overall growth rate (0.08 mm/year) observed in our study tracks closely with contemporary studies that have examined ascending aortic growth rates in association with normal aging and in cohorts with indolent aTAA.^10,15^ Interestingly, median baseline mid-AsAo diameter in the fastest growing Dramatic Growth group was 34 mm, which is both smaller than the median baseline size of the Stable group and at a level that would not be indicated for imaging surveillance in a patient of average body size. Similar findings have been noted in other studies that include patients with diameters below the conventional 40 mm cut-off, corroborating the idea that meaningful disease exists below current disease-defining thresholds.^15,16^ As expected,^5,10^ patients with Marfan syndrome or concomitant pathology in the distal aortic segments were more likely to be classified into growth groups that were more likely to experience TAAD, ascending aorta-specific mortality, and subsequent prophylactic ascending aortic repair – although effect sizes were modest.

A particularly interesting observation is that patients initially placed in a growth category often cross-over into a Stable or Non-Physiologic (i.e., noise-predominant) growth pattern – most likely reflecting the inherent intra- and inter-reader variability seen with manual diameter measurements, even under idealized conditions.^12^ For example, Non-Physiologic growth trajectories were ∼6 times more common at the “Extended” timepoint compared to the “Early” timepoint, suggesting that more surveillance doesn’t always add clarity to the patient’s growth trajectory. Notably, most patients initially classified as Stable remain so over surveillance, with rare (<0.3%) reclassification into the Dramatic Growth group associated with known risk-factors such as Marfan Syndrome (50%). Furthermore, the finding that patients classified as Dramatic Growth at the “Early” timepoint were just as likely to be classified as Non-Physiologic vs. remaining in Dramatic Growth after 5 scans mirrors a recent study finding that 50% of putative rapid ascending aorta ‘growers’ (3 mm/year) were found to be spurious on re-examination of the imaging.^14^ These findings imply that performing extended imaging surveillance, especially in stable disease, may be of limited or even negative value and should be avoided in the absence of clear and justified risk factors, as spurious values have the potential to complicate clinical interpretation and surgical management decisions.

### Parallels with oncological imaging

Current surveillance for ascending thoracic aortic aneurysm (aTAA) is challenged by the difficulty of distinguishing the few high-risk individuals from the vast majority with indolent disease, a problem which has many parallels with the issues of overdiagnosis and length-time bias in oncology screening.^16–18^ The fundamental challenge is that most dissections (80-90%) occur in patients with aortic diameters below the current surgical thresholds^8,9^ and only a small minority (∼16%) of patients who suffer an acute dissection were under surveillance for a known aneurysm beforehand.^19^

Borrowing from analogies in the oncologic literature, this suggests our current size-based strategies are well-suited to finding stable, slow-growing “tortoises,” but ill-equipped to identify the aggressive, fast-growing “hares” who would benefit most from intervention.^20,21^ Our findings support the applicability of this paradigm in ascending aortic disease and also highlight its potential harms. The LPA analysis revealed that most patients exhibit stable or very slow growth, and extended imaging often introduces more confusion than clarity. The prevalence of the “Non-physiologic” class—representing measurement noise—grew substantially from 1.3% in the “Early” analysis to 8.3% in the “Extended” analysis (p<0.001), suggesting more surveillance can amplify uncertainty. Such spurious findings do not show clear clinical benefit and can subject patients to anxiety and the risks of unnecessary interventions, including prophylactic surgery - an operation associated with an approximately 5% risk of major complications (i.e., stroke, myocardial infarction and/or death).^22^

### Clinical Implications

Our findings support de-escalating surveillance for the majority of patients whose ascending aortas are initially stable and are without evidence of other pathologies that predispose to repaid growth (e.g., aortitis, genetic aortopathy). In our cohort, nearly 80% of patients classified as ’Stable’ after 3 scans remained Stable with extended follow-up, and the transition to a ’Dramatic Growth’ class was rare (<1%). Crucially, this transition was almost exclusively observed in patients with established risk factors like Marfan syndrome. These results provide a data-driven rationale for reducing imaging frequency in stable patients who lack such high-risk features, which could alleviate patient anxiety and reduce healthcare system burden.

Conversely, our findings should prompt re-consideration of how we identify high-risk individuals. The fastest-growing “Dramatic Growth” group had significantly smaller baseline diameters than the Stable group (34 mm vs. 41 mm), and half of the patients who experienced a dissection had a baseline diameter of only 40 mm. Given that aortic dissection-related mortality has been increasing in several countries despite modern surveillance,^23^ focus may be shifted away from repeatedly monitoring stable disease in patients with mild to moderate dilation (i.e., 40-50 mm) and towards a more sophisticated search for the “hares.” This could be achieved by integrating comprehensive clinical assessment, such as a three-generation family history screening for aortic and vascular disease, clinical examinations for subtle signs of connective tissue disorders, and targeted genetic testing panels as recommended by current guidelines,^1^ to identify at-risk individuals who warrant intensive surveillance regardless of initial aortic size.

Emerging tools that leverage 3D shape and neural networks offer some potential paths for improvement in disease surveillance itself. U-Net approaches for automated and semi-automated segmentation of the aorta and landmark localization have shown promise for generating reliable diameter measurements from CT and MRI data,^24–26^ which may lessen the effects of variable and labor-intensive manual measurements. Further, techniques that leverage the entire 3D dataset to quantify aortic growth can help avoid the inherent challenges of quantifying a volumetric process (i.e., aortic growth) using a 1-dimensional diameter measurement.^27,28^ More reliable and precise ascending aortic measurements would enhance our ability to classify disease trajectory, possibly leading to better differentiation of risk at earlier time points or in patients with smaller absolute aortic size. Our findings also support calls for morphological features other than diameter (i.e., length^29^ or area^30^) and/or genetic markers^31^, to enhance our ability to identify and monitor high-risk patients.

### Limitations

Our study has several limitations. First, being a retrospective study there are factors that are difficult to accurately ascertain from medical records, such as detailed family history or patient or provider reasoning for lack of follow-up. Second, as a single-center study performed at an academic center with a dedicated aortic surveillance and surgery program, there is potential for referral bias. However, our patient demographics and TAAD event rates are similar to reports from other large surveillance cohorts.^5,7^ Third, there was only a small number of patients undergoing surveillance at sizes >50 mm in our study, as many of these patients undergo prophylactic repair above this threshold; interpretation of these results should be limited to pre-surgical populations undergoing surveillance. Lastly, the key outcomes of interest (TAAD, ascending aorta-specific mortality) are rare and so despite the inclusion of all eligible patients over nearly a twenty-year period, the event rates were low and more sophisticated statistical analyses of these outcomes was not possible.

## Conclusion

Our study demonstrates that the majority of patients undergoing imaging surveillance for aTAA exhibit indolent disease characterized by very slow or absent growth and a very low rate of acute complications, and that traditional methods of stratifying risk are insufficient. Given the variability/noise inherent to manual diameter measurements, extended imaging surveillance beyond 3 scans is unlikely to yield actionable clinical information or prevent TAAD, especially in those with stable disease and with an absence of known risk factors such as Marfan syndrome or aortitis. In addition, crossover between growth trajectory classes commonly occurs during surveillance, did not show clear association with key outcomes, and introduces the potential to promote waste and could result in harm. To improve aTAA management, a shift away from surveillance paradigms based on maximal ascending aortic diameter measurement appears warranted.

## Data Availability

Data for this study will be made available upon reasonable request and within the limitations of restrictions of sharing protected health information.

## Acknowledgments

We would like to acknowledge the Cardiovascular Health Improvement Project (CHIP) and MI-AORTA programs at the Frankel Cardiovascular Center for their support of this project through programmatic and database resources.

## Abbreviations

(aTAA): Ascending thoracic aortic aneurysm
(TAAD): Type A aortic dissection
(mid-AsAo): Mid-ascending aorta
(LPA): Latent Profile Analysis
(CTAs): CT-angiograms
(IQR): Interquartile range

## References

1. Isselbacher EM, Preventza O, Hamilton Black J, et al. 2022 ACC/AHA Guideline for the Diagnosis and Management of Aortic Disease: A Report of the American Heart Association/American College of Cardiology Joint Committee on Clinical Practice Guidelines. Circulation. 2022;146(24). doi:10.1161/cir.0000000000001106

2. Benedetti N, Hope MD. Prevalence and significance of incidentally noted dilation of the ascending aorta on routine chest computed tomography in older patients. J Comput Assist Tomogr. 2015;39(1):109–111. doi:10.1097/rct.0000000000000167

3. Mori M, Bin Mahmood SU, Yousef S, et al. Prevalence of Incidentally Identified Thoracic Aortic Dilations: Insights for Screening Criteria. Can J Cardiol. 2019;35(7):892–898. doi:10.1016/j.cjca.2019.03.023

4. Swahn E, Lekedal H, Engvall J, Nyström FH, Jonasson L. Prevalence and determinants of dilated ascending aorta in a Swedish population: a case-control study. Eur Heart J Open. 2023;3(5):oead085. doi:10.1093/ehjopen/oead085

5. Solomon MD, Leong T, Sung SH, et al. Association of Thoracic Aortic Aneurysm Size With Long-term Patient Outcomes: The KP-TAA Study. JAMA Cardiol. 2022;7(11):1160–1169. doi:10.1001/jamacardio.2022.3305

6. Wu J, Zafar MA, Liu Y, et al. Fate of the unoperated ascending thoracic aortic aneurysm: three-decade experience from the Aortic Institute at Yale University. Eur Heart J. 2023;44(43):4579–4588. doi:10.1093/eurheartj/ehad148

7. Kim JB, Spotnitz M, Lindsay ME, MacGillivray TE, Isselbacher EM, Sundt TM. Risk of Aortic Dissection in the Moderately Dilated Ascending Aorta. Journal of the American College of Cardiology. 2016;68(11):1209–1219. 10.1016/j.jacc.2016.06.025

8. Rylski B, Branchetti E, Bavaria JE, et al. Modeling of predissection aortic size in acute type A dissection: More than 90% fail to meet the guidelines for elective ascending replacement. J Thorac Cardiovasc Surg. 2014;148(3):944–8.e1. doi:10.1016/j.jtcvs.2014.05.050

9. Rylski B, Blanke P, Beyersdorf F, et al. How does the ascending aorta geometry change when it dissects? J Am Coll Cardiol. 2014;63(13):1311–1319. doi:10.1016/j.jacc.2013.12.028

10. Henry M, Campello Jorge CA, van Bakel PAJ, et al. Thoracic Aortic Aneurysm Growth Rates and Predicting Factors: A Systematic Review and Meta-Analysis. Journal of the American Heart Association. 2025;14(7):e038821. doi:10.1161/JAHA.124.038821

11. Marway PS, Tjahjadi N, Campello Jorge CA, et al. Baseline Diameter Does Not Predict Growth Rate in a Presurgical Ascending Thoracic Aortic Aneurysm Population. J Am Heart Assoc. 2024;13(20):e036896. doi:10.1161/JAHA.124.036896

12. Quint LE, Liu PS, Booher AM, Watcharotone K, Myles JD. Proximal thoracic aortic diameter measurements at CT: repeatability and reproducibility according to measurement method. Int J Cardiovasc Imaging. 2013;29(2):479–488. doi:10.1007/s10554-012-0102-9

13. Adriaans BP, Ramaekers M, Heuts S, et al. Determining the optimal interval for imaging surveillance of ascending aortic aneurysms. Neth Heart J. 2021;29(12):623–631. doi:10.1007/s12471-021-01564-9

14. Rapid Growth of Thoracic Aortic Aneurysm: Reality or Myth? Vol 167.; 2024. doi:10.1016/j.jtcvs.2022.06.021

15. Thijssen CGE, Mutluer FO, van der Toorn JE, et al. Longitudinal changes of thoracic aortic diameters in the general population aged 55 years or older. Heart. 2022;108(22):1767. doi:10.1136/heartjnl-2021-320574

16. Grimm LJ, Kruse DE, Tailor TD, Johnson KS, Allen BC, Ryser MD. Current Challenges in Imaging-Based Cancer Screening, From the AJR Special Series on Screening. American Journal of Roentgenology. Published online April 23, 2025. doi:10.2214/AJR.25.32808

17. Srivastava S, Koay EJ, Borowsky AD, et al. Cancer overdiagnosis: a biological challenge and clinical dilemma. Nat Rev Cancer. 2019;19(6):349–358. doi:10.1038/s41568-019-0142-8

18. Schieda N, Krishna S, Pedrosa I, Kaffenberger SD, Davenport MS, Silverman SG. Active Surveillance of Renal Masses: The Role of Radiology. Radiology. 2022;302(1):11–24. doi:10.1148/radiol.2021204227

19. Evangelista A, Isselbacher EM, Bossone E, et al. Insights From the International Registry of Acute Aortic Dissection. Circulation. 2018;137(17):1846–1860. doi:doi:10.1161/CIRCULATIONAHA.117.031264

20. Kattan MW. The Hypothetical Rabbit. Front Oncol. 2016;6:123. doi:10.3389/fonc.2016.00123

21. Hinman FJ. Screening for prostatic carcinoma. J Urol. 1991;145(1):126–129; discussion 129-130. doi:10.1016/s0022-5347(17)38267-8

22. Desai ND, Vekstein A, Grau-Sepulveda M, et al. Development of a Novel Society of Thoracic Surgeons Aortic Surgery Mortality and Morbidity Risk Model. The Annals of Thoracic Surgery. 2025;119(1):109–119. doi:10.1016/j.athoracsur.2024.09.025

23. Hibino M, Verma S, Jarret CM, et al. Temporal trends in mortality of aortic dissection and rupture in the UK, Japan, the USA and Canada. Heart. 2024;110(5):331–336. doi:10.1136/heartjnl-2023-323042

24. Katakol S, Baker TJ, Bian Z, et al. Fully automated pipeline for measurement of the thoracic aorta using joint segmentation and localization neural network. Journal of Medical Imaging. 2023;10(05). doi:10.1117/1.jmi.10.5.051810

25. Sedghi Gamechi Z, Bons LR, Giordano M, et al. Automated 3D segmentation and diameter measurement of the thoracic aorta on non-contrast enhanced CT. Eur Radiol. 2019;29(9):4613–4623. doi:10.1007/s00330-018-5931-z

26. Hamelink I (Iris), de Heide E (Erik J, Pelgrim GJ (Gert J, et al. Validation of an AI-based algorithm for measurement of the thoracic aortic diameter in low-dose chest CT. European Journal of Radiology. 2023;167:111067. doi:10.1016/j.ejrad.2023.111067

27. Burris NS, Bian Z, Dominic J, et al. Vascular Deformation Mapping for CT Surveillance of Thoracic Aortic Aneurysm Growth. Radiology. 2022;302(1):218–225. doi:10.1148/radiol.2021210658

28. Bian Z, Zhong J, Dominic J, Christensen GE, Hatt CR, Burris NS. Validation of a robust method for quantification of three-dimensional growth of the thoracic aorta using deformable image registration. Med Phys. 2022;49(4):2514–2530. doi:10.1002/mp.15496

29. Eagle Kim A., Bhave Nicole M. Ascending Aortic Length and Dissection Risk. JACC. 2019;74(15):1895–1896. doi:10.1016/j.jacc.2019.08.017

30. Acharya MN, Youssefi P, Soppa G, et al. Analysis of aortic area/height ratio in patients with thoracic aortic aneurysm and Type A dissection†. European Journal of Cardio-Thoracic Surgery. 2018;54(4):696–701. doi:10.1093/ejcts/ezy110

31. Ostberg NP, Zafar MA, Ziganshin BA, Elefteriades JA. The Genetics of Thoracic Aortic Aneurysms and Dissection: A Clinical Perspective. Biomolecules. 2020;10(2). doi:10.3390/biom10020182

